# The impact of competing risks in kidney allograft failure prediction

**DOI:** 10.1101/2024.05.13.24307280

**Authors:** Agathe Truchot, Marc Raynaud, Ilkka Helanterä, Olivier Aubert, Nassim Kamar, Christophe Legendre, Alexandre Hertig, Matthias Buchler, Marta Crespo, Enver Akalin, Gervasio Soler Pujol, Maria Cristina Ribeiro de Castro, Arthur J. Matas, Camilo Ulloa, Stanley C. Jordan, Edmund Huang, Ivana Juric, Nikolina Basic-Jukic, Maarten Coemans, Maarten Naesens, John J. Friedewald, Helio Tedesco Silva, Carmen Lefaucheur, Dorry L. Segev, Gary S. Collins, Alexandre Loupy

**Affiliations:** Université Paris Cité, INSERM U970 PARCC, Paris Institute for Transplantation and Organ Regeneration, F-75015 Paris, France; Department of Transplantation and Liver Surgery, Helsinki University Central Hospital, Helsinki, Finland; Kidney Transplant Department, Necker Hospital, Assistance Publique – Hôpitaux de Paris, Paris, France; Department of Nephrology and Organ Transplantation, Toulouse Rangueil University Hospital, INSERM UMR 1291, Toulouse Institute for Infectious and Inflammatory Diseases (Infinity), University Paul Sabatier, Toulouse, France; Department of Nephrology and Kidney Transplantation, Foch Hospital, Suresnes, France; Bretonneau Hospital, Nephrology and Immunology Department, Tours, France; Department of Nephrology, Hospital del Mar Barcelona, Spain; Albert Einstein College of Medicine, Renal Division Montefiore Medical Center, Kidney Transplantation Program, Bronx, NY, USA; Unidad de Trasplante Renopancreas, Centro de Educacion Medica e Investigaciones Clinicas Buenos Aires, Argentina; Hospital das Clinicas da Faculdade de Medicina da Universidade de Sao Paulo, Renal Transplantation Service Sao Paulo, Brazil; Division of Transplantation, Department of Surgery, University of Minnesota, Minneapolis, Minnesota, USA; Clinica Alemana de Santiago, Santiago, Chile; Department of Medicine, Division of Nephrology, Comprehensive Transplant Center, Cedars Sinai Medical Center, Los Angeles, CA, USA; Department of Nephrology, Arterial Hypertension, Dialysis and Transplantation, University Hospital Center Zagreb, School of Medicine University of Zagreb, Zagreb, Croatia; Department of Microbiology, Immunology and Transplantation, KU Leuven, Leuven, Belgium; Northwestern University Feinberg School of Medicine, Chicago, IL, USA; Universidade Federal de Sao Paulo, Hospital do Rim, Escola Paulista de Medicina, Sao Paulo, Brazil; Kidney Transplant Department, Saint-Louis Hospital, Assistance Publique - Hôpitaux de Paris, Paris, France; Department of Surgery, Johns Hopkins University School of Medicine, Baltimore, MD, USA; Centre for Statistics in Medicine, Nuffield Department of Orthopaedics, Rheumatology and Musculoskeletal Sciences, University of Oxford, Oxford, UK

**Keywords:** Cox, Fine-Gray, Kidney Transplantation, Graft Failure, Competing Risks

## Abstract

**Background:** Prognostic models are becoming increasingly relevant in clinical trials as potential surrogate endpoints, and for patient management as clinical decision support tools. However, the impact of competing risks on model performance remains poorly investigated. We aimed to carefully assess the performance of competing risks and non-competing risks models in the context of kidney transplantation, where allograft failure and death with a functioning graft are two competing outcomes.

**Methods:** We included 10 546 adult kidney transplant recipients enrolled in 10 countries (3941 patients in the derivation cohort, 6605 patients in international external validation cohorts). We developed prediction models for long-term kidney graft failure prediction, without accounting (i.e., censoring) and accounting for the competing risk of death with a functioning graft, using Cox and Fine-Gray regression models. To this aim, we followed a detailed and transparent analytical framework for competing and non-competing risks modelling, and carefully assessed the models’ development, stability, discrimination, calibration, overall fit, and generalizability in external validation cohorts and subpopulations. In total, 15 metrics were used to provide an exhaustive assessment of model performance.

**Results:** Among the 3941 recipients included in the derivation cohort, 538 (13.65%) lost their graft and 414 (10.50%) died after a median follow-up post-risk evaluation of 5.77 years (IQR 3.52-7.00). In the external validation cohorts, 896 (13.56%) graft losses and 525 (7.95%) deaths occurred after a median follow-up post-risk evaluation of 4.25 years (IQR 2.35-6.59). At 7 years post-risk evaluation, overestimation of the cumulative incidence was moderate when using Kaplan-Meier, compared to the Aalen-Johansen estimate (16.71% versus 15.67% in the derivation cohort). Cox and Fine-Gray models for predicting the long-term graft failure exhibited similar and stable risk estimates (average MAPE of 0.0140 and 0.0138 for Cox and Fine-Gray models, respectively). At 7 years post-risk evaluation, discrimination and overall fit were good and comparable in the external validation cohorts (concordance index ranging from 0.76 to 0.86, Brier Scores ranging from 0.102 to 0.141). In a large series of subpopulations and clinical scenarios, both models performed well and similarly.

**Conclusions:** Competing and non-competing risks models performed similarly in predicting long-term kidney graft failure. These results should be interpreted in light of the low rate of the competing event in our cohort, and do not stand as a general conclusion for competing risks modelling. Depending on the clinical scenario and the population considered, competing risks may be crucial to consider for accurate risk predictions.

## INTRODUCTION

Prognostic models have the potential to serve as companion tools for clinicians to enhance their prognostic judgements, optimize patient care, and personalize their follow-up^1^. They also become increasingly relevant in clinical trials as potential surrogate endpoints^2^. Models development studies need to be specifically designed towards the prediction of the clinical outcome. Not only are large, well-annotated, and deeply phenotyped cohorts required to capture a comprehensive set of candidate predictors^3^, but robust methodological standards for model development and validation must also be followed^4,5,6^. In many scenarios, this includes taking into account the presence of competing events, i.e., the medical event of interest being precluded by another earlier event, which is a frequently encountered setting when developing prognostic models^7^.

Handling competing events by censoring them assumes that the censoring mechanism is uninformative, implying that censored patients are expected to have the same likelihood of experiencing the event of interest as those who are not censored. Failing to account for competing risks can lead to an upward bias of the cumulative incidence of the event when estimated with the Kaplan-Meier estimator^8,9,10,11^. For instance, in studies with a recognized competing event – such as death in oncology when remission is the outcome^12,13^, or death in cardiology when non-fatal stroke is the outcome^14^ – the estimation of the cumulative incidence of the event of interest may be overestimated when censoring for the competing event.

In kidney transplantation, prediction of allograft failure is crucial for optimal patient management. This outcome is generally censored for death, that is, for patients who die with a functioning allograft, it is generally assumed that the allograft is still functioning at the time of death. This assumption has been recently criticized because of the resulting overestimation of the cumulative incidence of allograft failure^15^. One strategy would consist in regrouping allograft failure and death into a composite endpoint (e.g. “all-cause graft loss”). Nevertheless, this approach assumes that the competing event shares the same set of predictors that influence both events in the same way^16,17^.

Conversely, keeping distinct outcomes and accounting for the competing risk of death should be considered^18,19,20^. However, blindly applying competing risks modelling without considering issues related to the design and the aim of the study might also be inappropriate. Moreover, the impact of accounting for the competing event on prediction model performance and generalizability is unclear. Despite current recommendations^21,22,23^, there is a dearth of research of assessing the impact of competing risks on model performance.

Therefore, we aimed to investigate the impact of competing risks on long-term kidney graft failure prediction. To this aim, we used large, international, deeply phenotyped cohorts of kidney recipients and assessed whether the development and validation of a prognostic model without accounting for the competing risk of death were affected by this competing event.

## METHODS

### Study design and participating cohorts

We included kidney transplant recipients from the Paris Transplant Group qualified database^24^ (NCT03474003) aged 18 years and older prospectively enrolled on the day of transplantation, either from a living or deceased donor, in France between 1st January 2005 and 1st January 2014. Patients experiencing allograft primary non-function were excluded from the analyses. Clinical data were collected from each centre and entered into the Paris Transplant Group database system (French Data Protection Authority registration no. 363505), using a structured protocol to ensure harmonization across study centres. All data were anonymized and prospectively entered at the time of transplantation, at the time of post-transplant allograft biopsies, and at each transplant anniversary using a standardized protocol. Allograft outcomes were prospectively assessed until January 1, 2021. To ensure data accuracy, an annual audit was performed.

External validation was carried out on multiple international datasets of kidney transplant recipients (living or deceased donation), involving 23 centres in 10 countries: France (n=1733), Belgium (n=838), Spain (n=133), Croatia (n=314), Finland (n=413), United States and Canada (n=2384), Argentina (n=135), Brazil (n=530), and Chile (n=125). Data were collected and entered in the databases of the centres in accordance with local and national regulatory standards, and submitted to the Paris Transplant Group anonymously. In each cohort, patients gave written informed consent on the day of transplantation.

A total of 10 546 kidney transplant recipients were included for the final analyses, this included 3941 in the derivation cohort and 6605 in the external validation cohorts. External validation cohorts’ data were combined into a European validation cohort (European transplant centres, n=3431), a North American validation cohort (US and Canadian transplant centres, n=2384), and a South American validation cohort (South American transplant centres, n=790).

### Data collection and procedures

To investigate the key prognostic determinants of allograft outcome, patients were extensively phenotyped, encompassing donor and recipient-related demographic characteristics, transplant characteristics, biological parameters, as well as immunological and histological parameters. The following candidate predictors were collected: (1) recipient and donor characteristics including age, sex, and comorbidities; (2) donor characteristics including age, sex, serum creatinine, deceased or living, cause of death, history of hypertension or diabetes; (3) transplant characteristics including previous kidney transplant, cold ischemia time, number of HLA mismatches, delayed graft function; (4) functional parameters including estimated glomerular filtration rate (eGFR) by the Modification of Diet in Renal Disease Study equation, and the proteinuria level using the urine protein/creatinine ratio; (5) immunological parameters including circulating anti-human leukocyte antigen donor-specific antibody specificities and mean fluorescence intensity specificities and levels; and (6) allograft histopathology data including glomerulitis, transplant glomerulopathy, tubulitis, interstitial inflammation, interstitial fibrosis and tubular atrophy, endarteritis, arteriosclerosis, arteriolar hyalinosis, peritubular capillaritis and C4d deposition (g, cg, t, i, IFTA, v, cv, ah, ptc, and C4d Banff scores), and diagnoses.

We defined the time of initial risk evaluation as the time of allograft biopsy after transplantation. Kidney transplant biopsies were performed as per protocol and for clinical indication. At the time of risk evaluation, recipients underwent concomitant evaluation of eGFR and proteinuria, allograft biopsy (Banff lesion scores and diagnoses), and circulating anti-HLA antibody. In the external validation cohorts, in case of multiple biopsies, the closest to one year post-transplantation was chosen.

### Outcome measures

Death-censored allograft survival was the outcome that we aimed to predict when ignoring the competing risk of death, and non-death-censored allograft survival was the outcome that we aimed to predict when accounting for the competing risk of death. In the case of death-censored allograft survival, patients who died with a functioning allograft were censored at the time of death. Allograft failure was defined as a patient’s definitive return to dialysis or pre-emptive kidney re-transplantation.

Follow-up started from the patient’s initial risk evaluation up to the date of allograft failure, death, or the end of the follow-up (01/01/2021). The maximum follow-up was truncated to 7 years.

### Statistical Analysis

Continuous variables were described using means and standard deviations (SDs) or medians and interquartile ranges, as appropriate. Means and proportions between groups were compared with Student’s t-test, analysis of variance (ANOVA) or the chi-square test (or Fisher’s exact test if appropriate). Values of *P* < 0.05 were considered significant, and all tests were two-tailed.

#### Descriptive survival analysis

In the case of death-censored graft survival, graft survival was estimated with the Kaplan-Meier estimator^25^. When accounting for the competing risk of death, graft survival was estimated with the Aalen-Johansen estimator^26,27^.

#### Competing and non-competing risks modelling frameworks

##### Prediction model development

In the derivation cohort (Paris Transplant Group database), a multivariable Cox model was used to predict long-term death-censored graft failure, integrating eight independent clinically relevant parameters derived from the large set of candidate factors: 1) kidney-graft function assessed by the eGFR and proteinuria level, 2) circulating donor-specific antibodies, 3) kidney-graft pathology data with transplant glomerulopathy (cg Banff score), microcirculation inflammation (g+ptc Banff score), interstitial fibrosis and tubular atrophy (IFTA Banff score) and interstitial inflammation and tubulitis (i+t Banff score) recorded according to the Banff classification and 4) the delay between the date of transplantation and the date of risk evaluation. This model (the iBox score^28^) is specifically designed towards long-term death-censored graft failure and stands as the most validated model so far in kidney transplantation^29,30^. It has been qualified as a secondary endpoint for clinical trials by the European Medicine Agency^24^.

To account for the competing risk of death in long-term graft failure prediction, a Fine-Gray subdistribution hazards model^31^ integrating the independent parameters listed above was developed on the derivation cohort. The Fine-Gray model was chosen for its ability to directly estimate the cumulative incidence of the outcome of interest in the presence of competing risks, thus facilitating the evaluation of predictive accuracy and comparison.

##### Prediction model stability

Model stability was investigated by producing 500 bootstrap models and deriving prediction instability plots and instability index, as described in Riley and Collins^32^. Selection of the predictors was not replicated in the bootstrap samples. Prediction instability plots were obtained by plotting bootstrap predictions against original predictions. Mean absolute prediction error (MAPE) values were calculated for each observation and MAPE instability plots were obtained by plotting individual MAPE values against original predictions.

##### Evaluation of model performance

Performance of the Cox and Fine-Gray models were assessed in terms of discrimination, calibration, and overall fit in the derivation cohort and in the three external validation cohorts. The chosen time horizon was 7 years post-risk evaluation in the derivation cohort and in the European and North American validation cohorts, and 5 years post-risk evaluation in the South American cohort, due to the shorter follow-up in this cohort.

First, the discrimination was assessed with Harrell’s concordance index (Harrell’s c-index)^33^, and Uno’s concordance index (Uno’s c-index)^34^. Respectively for Cox and Fine-Gray model, the expected mortality^35^, and the cause-j mortality^36^ were used as one-dimensional summaries of relative risk predictions for concordance evaluation^37^.

Second, calibration was assessed in terms of calibration slope, calibration-in-the-large, and observed/expected ratio. For the Cox model, the calibration slope was obtained by regressing the prognostic index (PI) values of the Cox model with the difference between the cumulative hazard (log transformation) and the PI as an offset using a Poisson model, and calibration intercept was obtained by regressing the cumulative hazard (log transformation) as an offset using a Poisson model^38^. For the Fine-Gray model, the calibration slope was estimated by regressing the pseudo-observations with the risk estimates (log-log transformation) as an offset using a generalized linear model^39^. The observed/expected ratio was obtained by dividing the observed outcome proportion given by the Kaplan-Meier estimator (for the Cox model) or the Aalen-Johansen estimator (for the Fine-Gray model) and the expected risk given by the mean of (one minus) the predicted survival probabilities (for the Cox model) and the mean of the predicted risks (for the Fine-Gray model).

Calibration was also assessed graphically. To produce calibration plots, predicted risks from the Cox and Fine-Gray models were divided into equally-sized groups and, for each group, the median was plotted against the observed event probability estimated by (one minus) the Kaplan-Meier estimator for the Cox model and the Aalen-Johansen estimator for the Fine-Gray model, respectively. In each cohort, the number of groups was chosen to ensure a minimum of 100 observations per group comprising, if possible, at least 10 events (10 graft losses or 10 graft losses and 10 deaths). Smoothed calibration curves were obtained by two approaches: first, by fitting a secondary Cox (respectively, Fine-Gray) model with restricted cubic splines (three knots) to the risk estimates (log-log transformation) obtained from the Cox (Fine-Gray) prediction models, as described by Austin *et al* for non-competing risks^40^ and competing risks^41^; second, by deriving pseudo-observations^39^ and using loess smoothing with a span of 0.75 through the risk estimates and these pseudo-observations.

Additionally, the Integrated Calibration Index (ICI) and its quantiles E50, E90 and Emax and the root squared bias were obtained using the aforementioned restricted cubic splines. These metrics allow for a comparison of the relative calibration of different prediction models. Third, the overall fit was assessed with the Brier Score and its derivatives (the Integrated Brier Score and the Index of Predictive Accuracy^42^), which capture both calibration and discrimination, and Royston and Sauerbrei’s R^2^_D_^43^. The Brier Scores were calculated using inverse probability of censoring weighting, as defined by Gerds *et al*^44^ for non-competing risks and by Schoop *et al*^45^ for competing risks. The Integrated Brier Score was calculated as the integration of the Brier Score over the time range of interest (0 to 7 years or 0 to 5 years). The Index of Predictive Accuracy is a scaled version of the Brier Score, defined as one minus the Brier Score of the prediction model divided by the Brier Score of the “null model”. The Brier Score for the “null model” is obtained with the Kaplan-Meier estimator for the non-competing risks framework and with the Aalen-Johansen estimator for the competing risks framework.

##### Confidence intervals

For the Cox model, 95% confidence intervals for concordance metrics (Harrell’s c-index and Uno’s c-index) were based on Therneau et Atkinson^46^. For Cox and Fine-Gray models, calibration slope and intercept are presented along normal-based 95% confidence intervals, and Brier Scores’ 95% confidence intervals were based on Blanche *et al*^47^. For all the other metrics, for both models, confidence intervals were obtained using bootstrap on 500 samples, with the 2.5th and 97.5th percentile as values for the lower and upper bounds.

#### Internal validation

In the derivation cohort, optimism-corrected performance for Cox and Fine-Gray models were obtained by randomly bootstrapping the data 500 times and calculating the average difference between the model performance in the original data and in the bootstrap samples after fitting the models in each bootstrap sample.

#### Missing data

There were no missing values for follow-up time, outcome status and predictors included in both models in any of the derivation or validation cohorts.

#### Subgroup analyses

Subgroup analyses were conducted to evaluate the robustness and magnitude of performance divergence in both models across various subpopulations and clinical scenarios within the derivation cohort. We stratified patients by age, sex, BMI, race, type of treatment induction, immunological risk, deceased or living donors, older donors, expanded criteria donors, and we implemented different timings of risk evaluation.

#### Software

All analyses were performed using R (version 4.0.4, R Foundation for Statistical Computing, Vienna, Austria).

## RESULTS

### Cohorts’ characteristics

Overall, 10 546 patients from 10 countries were included in the study. The derivation cohort included a total of 3941 patients from four centres, and the external validation cohorts included a total of 6605 patients from 19 centres. The median time from transplantation to risk evaluation was 0.98 years (IQR 0.27-1.07) in the derivation cohort and 1.00 years (IQR 0.46-1.13) in the validation cohorts. Restricting to 7-years post risk-evaluation, 538 (13.65%) graft losses and 414 (10.50%) deaths occurred in the derivation cohort after a median follow-up post-risk evaluation of 5.77 years (IQR 3.52-7.00), and 896 (13.56%) graft losses and 525 (7.95%) deaths occurred in the validation cohorts after a median follow-up post-risk evaluation of 4.25 years (IQR 2.35-6.59). Characteristics of the derivation and validation cohorts are shown in table 1.

**Table 1:** Characteristics of the development and validation cohorts (n=10 546)

|  | French<br>derivation cohort<br>N=3941 |  | European<br>Validation cohort<br>N=3431 |  | North American<br>validation cohort<br>N=2384 |  | South American<br>validation cohort<br>N=790 |  | P |
| --- | --- | --- | --- | --- | --- | --- | --- | --- | --- |
|  | n |  | n |  | n |  | n |  |  |
| Recipient characteristics |  |  |  |  |  |  |  |  |  |
| Age (years) mean (SD) | 3941 | 49.8 (13.7) | 3431 | 51.4 (13.6) | 2384 | 49.4 (13.9) | 790 | 43.3 (15.4) | <0.001 |
| Gender male No. (%) | 3941 | 2416 (61.3%) | 3431 | 2170 (63.2%) | 2374 | 1436 (60.5%) | 790 | 483 (61.1%) | 0.150 |
| Cause of end stage renal disease | 3941 |  | 2821 |  | 1534 |  | 721 |  | <0.001 |
| Glomerulonephritis No. (%) |  | 1070 (27.2%) |  | 818 (29.0%) |  | 380 (24.8%) |  | 178 (24.7%) |  |
| Diabetes No. (%) |  | 432 (11.0%) |  | 394 (14.0%) |  | 386 (25.2%) |  | 80 (11.1%) |  |
| Vascular No. (%) |  | 291 (7.38%) |  | 220 (7.80%) |  | 255 (16.6%) |  | 128 (17.8%) |  |
| Other No. (%) |  | 2148 (54.5%) |  | 1389 (49.2%) |  | 513 (33.4%) |  | 335 (46.5%) |  |
| Donor characteristics |  |  |  |  |  |  |  |  |  |
| Age (years) mean (SD) | 3941 | 51.6 (16.3) | 3424 | 50.1 (15.5) | 2375 | 40.8 (14.5) | 786 | 45.0 (14.7) | <0.001 |
| Male gender No. (%) | 3941 | 2123 (53.9%) | 2842 | 1615 (56.8%) | 2382 | 1159 (48.7%) | 787 | 414 (52.6%) | <0.001 |
| Hypertension No. (%) | 3847 | 990 (25.7%) | 2070 | 543 (26.2%) | 1654 | 230 (13.9%) | NA | NA* | <0.001 |
| Diabetes mellitus No. (%) | 3806 | 228 (5.99%) | 1096 | 64 (5.84%) | 1651 | 73 (4.42%) | NA | NA* | 0.062 |
| Donor type |  |  |  |  |  |  |  |  |  |
| Deceased donor No. (%) | 3941 | 3279 (83.2%) | 2995 | 2718 (90.8%) | 2384 | 1216 (51.0%) | 788 | 543 (68.9%) | <0.001 |
| Death from cerebrovascular disease No. (%) | 3941 | 1837 (46.6%) | 3006 | 922 (30.7%) | 1168 | 266 (22.8%) | NA | NA* | <0.001 |
| Expanded criteria donor No. (%) | 3936 | 1387 (35.2%) | 1553 | 507 (32.6%) | 1409 | 212 (15.0%) | 746 | 182 (24.4%) | <0.001 |
| Transplant characteristics |  |  |  |  |  |  |  |  |  |
| Prior kidney transplant No. (%) | 3941 | 596 (15.1%) | 3393 | 454 (13.4%) | 1286 | 215 (16.7%) | 782 | 83 (10.6%) | <0.001 |
| Cold ischemia time in deceased donors (hours) mean (SD) | 3917 | 16.2 (8.99) | 2393 | 15.2 (6.85) | 1708 | 9.98 (11.0) | 758 | 17.1 (10.6) | <0.001 |
| HLA-A/B/DR mismatch number mean (SD) | 3941 | 3.82 (1.36) | 3392 | 3.19 (1.42) | 1503 | 3.67 (1.75) | 656 | 2.71 (1.37) | <0.001 |
| Delayed graft function No. (%) | 3841 | 1035 (26.9%) | 3273 | 671 (20.5%) | 2287 | 312 (13.6%) | 734 | 340 (46.3%) | <0.001 |
| Time from transplantation to risk evaluation (years) median (IQR) | 3941 | 0.98 [0.27;1.07] | 3431 | 1.00 [0.28;1.04] | 2384 | 1.00 [0.53;1.12] | 790 | 1.96 [0.96;3.47] | <0.001 |
| Functional parameters at time of risk evaluation |  |  |  |  |  |  |  |  |  |
| eGFR (mL/min/1.73 m<sup>2</sup>) mean (SD) | 3941 | 49.8 (19.4) | 3431 | 50.5 (20.9) | 2384 | 48.9 (22.6) | 790 | 38.4 (18.8) | <0.001 |
| Proteinuria (g/g) median (IQR) | 3941 | 0.19 (0.10 - 0.39) | 3431 | 0.15 [0.09;0.35] | 2384 | 0.19 [0.05;1.14] | 790 | 0.27 [0.05;0.75] | <0.001 |
| Immunological parameters at time of risk evaluation |  |  |  |  |  |  |  |  |  |
| Anti-HLA donor specific antibody mean fluorescence intensity | 3941 |  | 3431 |  | 2384 |  | 790 |  | <0.001 |
| < 1400 No. (%) |  | 3607 (91.5%) |  | 3217 (93.8%) |  | 2064 (86.6%) |  | 714 (90.4%) |  |
| ≥ 1400 No. (%) |  | 334 (8.48%) |  | 214 (6.24%) |  | 320 (13.4%) |  | 76 (9.62%) |  |
| Outcomes at 7 years post-risk evaluation |  |  |  |  |  |  |  |  |  |
| Graft loss No. (%) | 3941 | 538 (13.7%) | 3431 | 410 (11.9%) | 2384 | 338 (14.2%) | 790 | 148 (18.7%) | <0.001 |
| Death No. (%) | 3941 | 414 (10.5%) | 3431 | 363 (10.6%) | 2384 | 117 (4.91%) | 790 | 45 (5.70%) | <0.001 |

eGFR, estimated glomerular filtration rate; HLA, human leukocyte antigen; IQR, interquartile range; NA, not available. \*Data not available for the South American cohort, P value refers to the 3 other cohorts.

### Estimation of the long-term allograft survival with Kaplan-Meier and Aalen-Johansen

Cumulative incidences of graft loss with and without accounting for the competing risk of death are shown in figure 1 and in supplementary table 1, in the derivation and validation cohorts. The Kaplan-Meier (KM) estimations of cumulative incidences were greater than the corresponding Aalen-Johansen (AJ) estimations. This overestimation was however moderate, with 1-KM and AJ estimates of 16.71% and 15.67% at 7 years, respectively, in the derivation cohort (relative difference of 6.63%), 16.11% and 15.01% in the European validation cohort (relative difference of 7.31%), 22.12% and 21.25% in the North American validation cohort (relative difference of 4.07%), and 36.92% and 34.04% in the South American validation cohort (relative difference of 8.48%).

**Figure 1.**
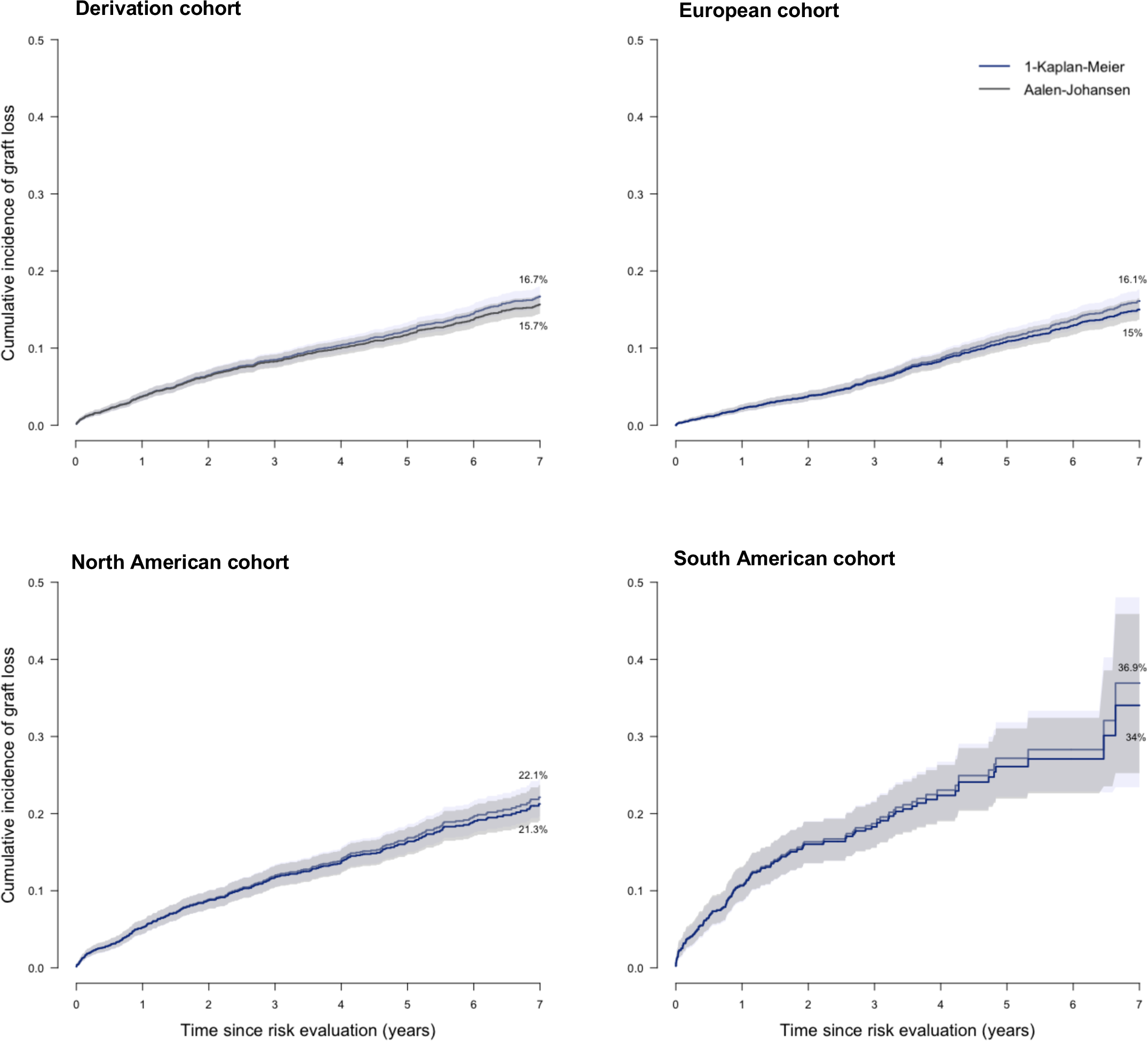
Cumulative incidence functions up to 7 years post risk-evaluation in the derivation and validation cohorts. Cumulative incidence of graft loss when ignoring the competing risk of death was estimated with the Kaplan-Meier estimator, and with the Aalen-Johansen estimator when accounting for the competing risk of death, in the derivation and external validation cohorts, ranging from 0 to 7 years post-risk evaluation.

### Development of the Cox and Fine-Gray models to predict the long-term allograft survival

The coefficients of the determinants of graft loss, as estimated by Cox and Fine-Gray models, are shown in figure 2 and supplementary table 2. The estimated effects of proteinuria, IFTA Banff score and cg Banff score (subHR_proteinuria_=1.44, subHR_IFTA=3_ =1.37 and subHR_cg≥1_= 1.32) were smaller than their corresponding effects on the rate (HR_proteinuria_=1.50, HR_IFTA=3_ =1.41 and HR_cg≥1_= 1.41). Time from transplant to evaluation (subHR 1.07 [1.00 – 1.14]) and cg Banff score (subHR 1.32 [CI 1.00-1.75]) were slightly out of significance in the Fine-Gray model.

**Figure 2.**
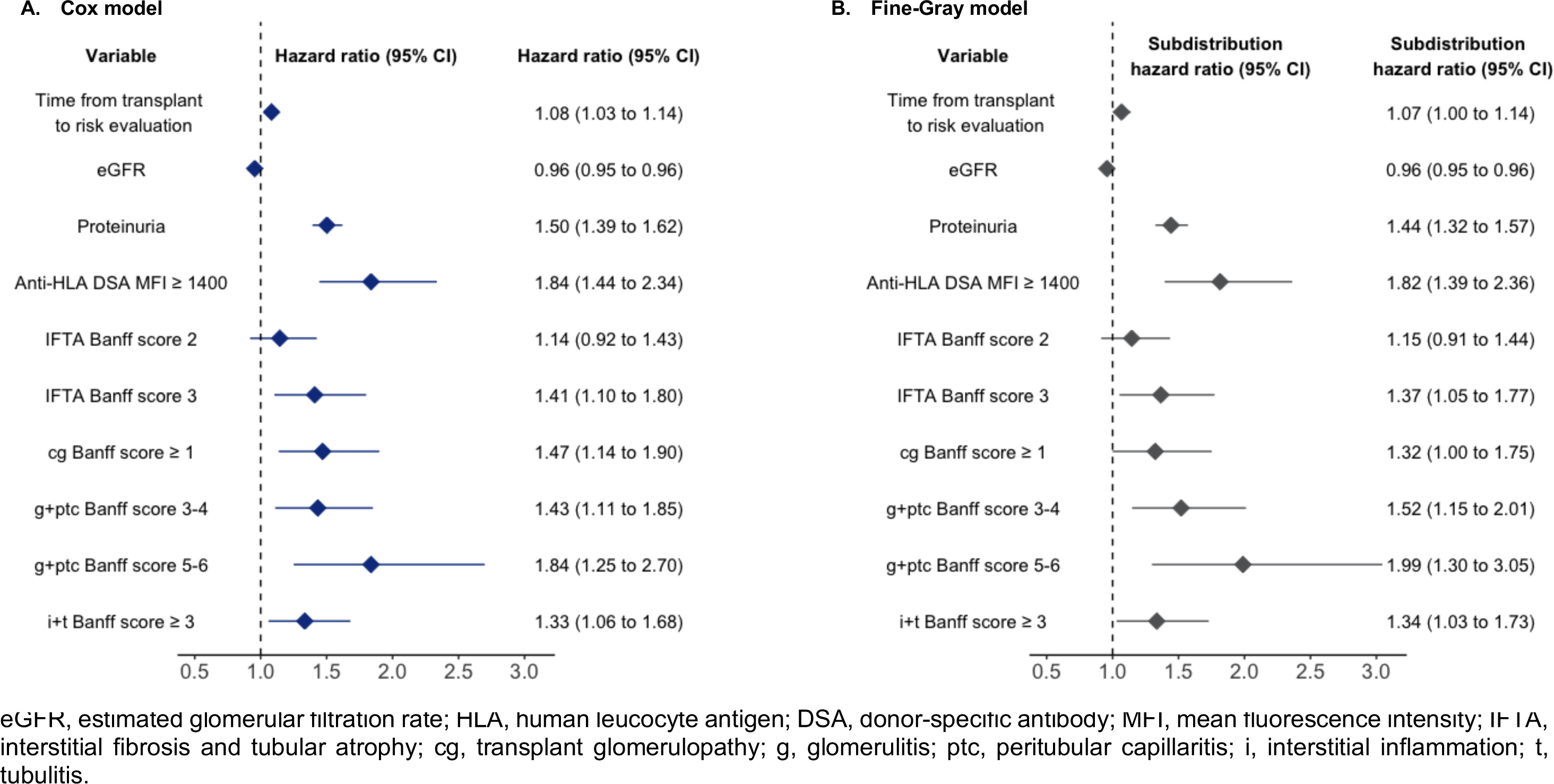
Cox and Fine-Gray multivariable models. Models are presented as their exponentiated coefficients (hazard ratios and subdistribution hazard ratios with 95% confidence intervals) for the eight independent determinants of kidney allograft loss assessed at time of post-transplant risk evaluation in the derivation cohort. The Cox model is presented in the left panel (A) and the Fine-Gray model in the right panel (B).

Comparison of 7-years predictions from the Cox and Fine-Gray model are shown in figure 3, along with a loess curve and the P20 (the proportion of the Fine-Gray’s predictions within 20% of the Cox’s predictions). For all the cohorts, 99% of Fine-Gray model’s predictions fell within 20% of the Cox model’s predictions.

**Figure 3.**
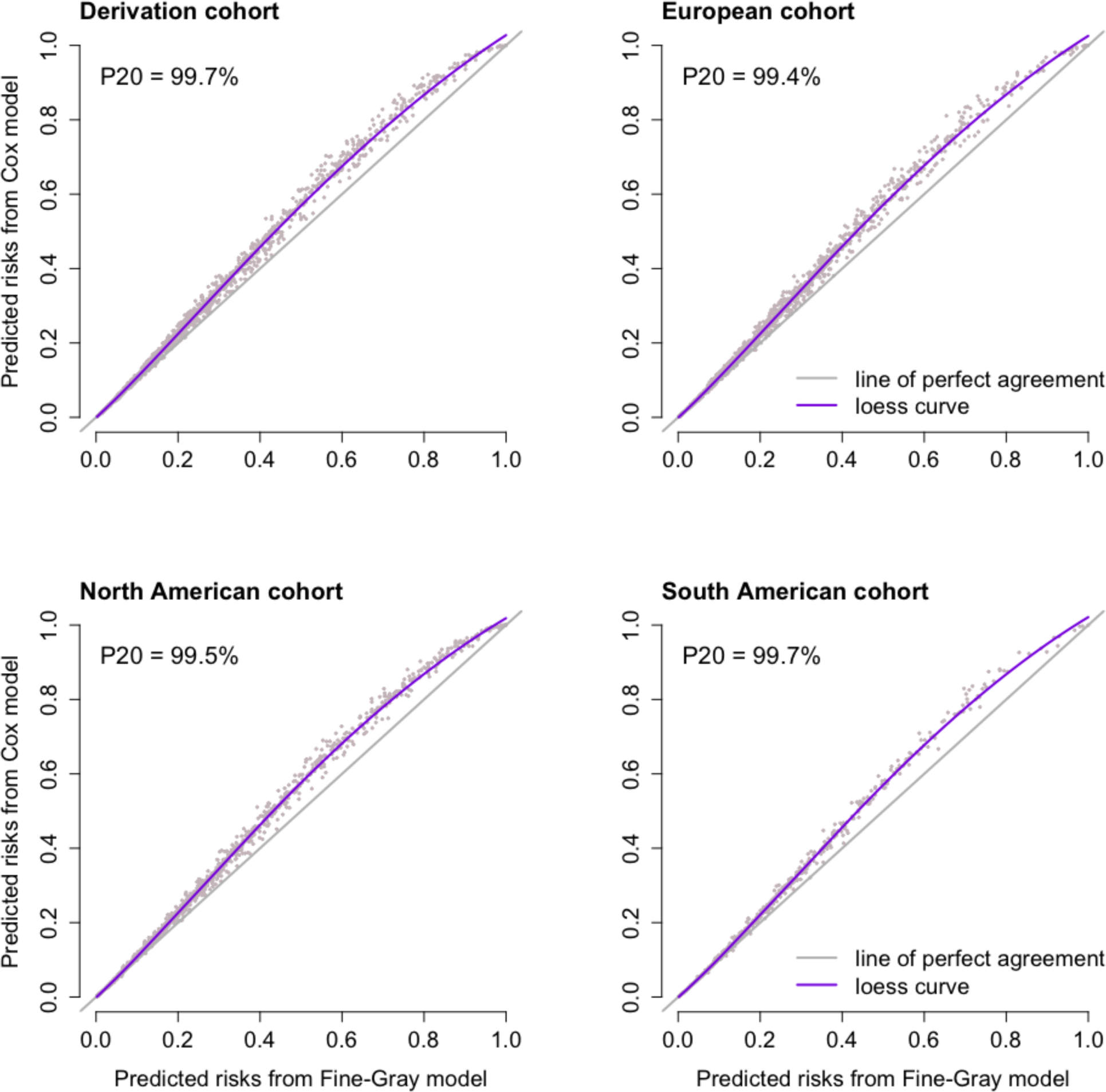
Fine-Gray model’s predictions against Cox model’s predictions at 7 years post-risk evaluation. Distribution of Fine-Gray model’s predictions according to Cox model’s predictions in the derivation and external validation cohorts. Cox model’s predictions refer to one minus the individual predicted survival probabilities at 7 years post risk-evaluation. Fine-Gray model’s predictions refer to the individual predicted risks (cumulative incidence) at 7 years post risk-evaluation. P20 is the proportion of Fine-Gray model’s predictions within 20% of Cox model’s predictions.

Prediction instability plots and MAPE instability plots for both models are shown in supplementary figure 1. Cox and Fine-Gray models exhibited close and stable individual risk estimates, with an average MAPE of 0.0140 and 0.0138, respectively.

### Predictive performance of the Cox and Fine-Gray models

Discrimination, calibration and overall fit are shown in table 2 for the different cohorts.

**Table 2.** Predictive performance of the Cox and Fine-Gray models at 7 years post-risk evaluation. Discrimination, calibration and overall accuracy performance metrics for the Cox and Fine-Gray models assessed at 7 years post risk-evaluation in the derivation cohort and in the European and North American external validation cohorts, and at 5 years post-risk evaluation in the South American external validation cohort. Performance metrics are presented along with 95% confidence intervals, except for the optimism-corrected performance of the derivation cohort

| Performance metrics:<br>7 years post-evaluation | Apparent performance |  | Internal validation: optimism-corrected performance<br>B=500 bootstraps |  | External validation |  |  |  |  |  |
| --- | --- | --- | --- | --- | --- | --- | --- | --- | --- | --- |
|  | Derivation cohort<br>n=3,941 |  | Derivation cohort<br>n=3,941 |  | European cohort<br>n=3,431 |  | North American cohort<br>n=2,384 |  | South American cohort*<br>n=790 |  |
|  | Cox | Fine-Gray | Cox | Fine-Gray | Cox | Fine-Gray | Cox | Fine-Gray | Cox | Fine-Gray |
| Discrimination |  |  |  |  |  |  |  |  |  |  |
| Harrell's c-index | 0.809<br>[0.790;0.827] | 0.800<br>[0.781;0.817] | 0.807 | 0.797 | 0.770<br>[0.747;0.792] | 0.762<br>[0.739;0.783] | 0.814<br>[0.789;0.836] | 0.805<br>[0.784;0.828] | 0.862<br>[0.831;0.888] | 0.855<br>[0.825;0.882] |
| Uno's c-index | 0.791<br>[0.769;0.810] | 0.785<br>[0.765;0.804] | 0.788 | 0.782 | 0.758<br>[0.732;0.781] | 0.752<br>[0.726;0.776] | 0.781<br>[0.751;0.808] | 0.774<br>[0.748;0.801] | 0.817<br>[0.773;0.855] | 0.812<br>[0.774;0.847] |
| Calibration |  |  |  |  |  |  |  |  |  |  |
| Slope | 1<br>[0.931;1.068] | 0.900<br>[0.811;0.990] | 0.981 | 0.882 | 0.696<br>[0.625;0.766] | 0.913<br>[0.777;1.049] | 0.847<br>[0.767;0.928] | 0.785<br>[0.650;0.920] | 1.200<br>[1.048;1.352] | 1.056<br>[0.773;1.338] |
| Intercept | 0<br>[-0.086;0.083] | -0.057<br>[-0.157;0.044] | -0.010 | -0.064 | -0.303<br>[-0.402;-0.207] | -0.073<br>[-0.194;0.048] | 0.057<br>[-0.053;0.163] | -0.034<br>[-0.179;0.111] | 0.388<br>[0.218;0.549] | 0.364<br>[0.152;0.577] |
| O/E ratio | 0.935<br>[0.871;1.003] | 0.970<br>[0.902;1.042] | 0.934 | 0.970 | 0.939<br>[0.854;1.020] | 0.972<br>[0.886;1.054] | 0.967<br>[0.870;1.066] | 1.034<br>[0.928;1.138] | 1.186<br>[0.994;1.415] | 1.259<br>[1.060;1.493] |
| Overall fit |  |  |  |  |  |  |  |  |  |  |
| Brier Score | 0.102<br>[0.094;0.111] | 0.103<br>[0.095;0.111] | 0.103 | 0.104 | 0.120<br>[0.109;0.130] | 0.114<br>[0.104;0.123] | 0.115<br>[0.099;0.131] | 0.120<br>[0.105;0.135] | 0.139<br>[0.106;0.172] | 0.141<br>[0.111;0.171] |
| Integrated Brier Score | 0.054<br>[0.049;0.059] | 0.054<br>[0.050;0.059] | 0.054 | 0.055 | 0.057<br>[0.051;0.062] | 0.054<br>[0.050;0.059] | 0.069<br>[0.062;0.076] | 0.070<br>[0.063;0.077] | 0.076<br>[0.064;0.088] | 0.079<br>[0.067;0.090] |
| IPA, % | 26.4<br>[22.5;29.8] | 22.0<br>[18.4;25.2] | 25.6 | 21.2 | 11.4<br>[5.1;17.2] | 10.9<br>[5.7;15.7] | 33.4<br>[27.9;39.2] | 28.4<br>[23.1;33.4] | 29.8<br>[17.2;41.3] | 27.0<br>[16.3;36.2] |
| Royston R<sup>2</sup><sub>D</sub> | 0.520<br>[0.477;0.561] | 0.487<br>[0.441;0.532] | 0.510 | 0.475 | 0.341<br>[0.295;0.386] | 0.342<br>[0.294;0.385] | 0.518<br>[0.469;0.571] | 0.496<br>[0.445;0.549] | 0.585<br>[0.511;0.650] | 0.586<br>[0.509;0.643] |

O/E ratio: observed/expected ratio; IPA, index of predictive accuracy \*Performance metrics were assessed at 5 years post-risk evaluation in the South American cohort.

#### Discrimination

Both models showed good and comparable discrimination in the derivation and validation cohorts (Harrell’s c-index range 0.76 to 0.86), at 7 years post-risk evaluation. In the derivation cohort, the Cox model achieved a c-index of 0.809 [CI 0.790;0.827] (optimism-corrected performance 0.807), and 0.800 [CI 0.781;0.817] (optimism-corrected performance 0.797) for the Fine-Gray model. In the validation cohorts, the c-index values were similar between the Cox model and the competing risk model: 0.770 [CI 0.747;0.792] and 0.762 [CI 0.739;0.783] in the European cohort, 0.814 [CI 0.789;0.836] and 0.805 [CI 0.784;0.828] in the North American cohort and 0.862 [CI 0.831;0.888] and 0.855 [CI 0.825;0.882] in the South American cohort. Correction by inverse probability of censoring weighting resulted in decreased but good discriminative estimates (Uno’s c-index range 0.75 to 0.81) for both models.

#### Calibration

Both Cox and Fine-Gray models showed close and good agreement between the predicted and observed risks in the derivation cohort and in the North American cohort (figure 4 and supplementary figure 2, table 2). In the European validation cohort, the models tended to overestimate the risks, as reflected in the negative calibration intercept (−0.303 [CI −0.402;-0.207] and −0.073 [CI −0.194;-0.048] for Cox and Fine-Gray, respectively) and the O/E ratio lower than 1 (0.939 [CI 0.854;1.020] and 0.972 [CI 0.886;1.054] for Cox and Fine-Gray, respectively). In this cohort, the Fine-Gray model was slightly better calibrated than the Cox model (figure 4, table 2). In the South American validation cohort, both models tended to underestimate the risks (positive calibration intercept 0.388 [CI 0.218;0.549] and 0.364 [CI 0.152;0.577] for Cox and Fine-Gray, respectively), and observed/expected ratio greater than one (1.186 [CI 0.994;1.415] and 1.259 [CI 1.060;1.493] for Cox and Fine-Gray, respectively).

**Figure 4.**
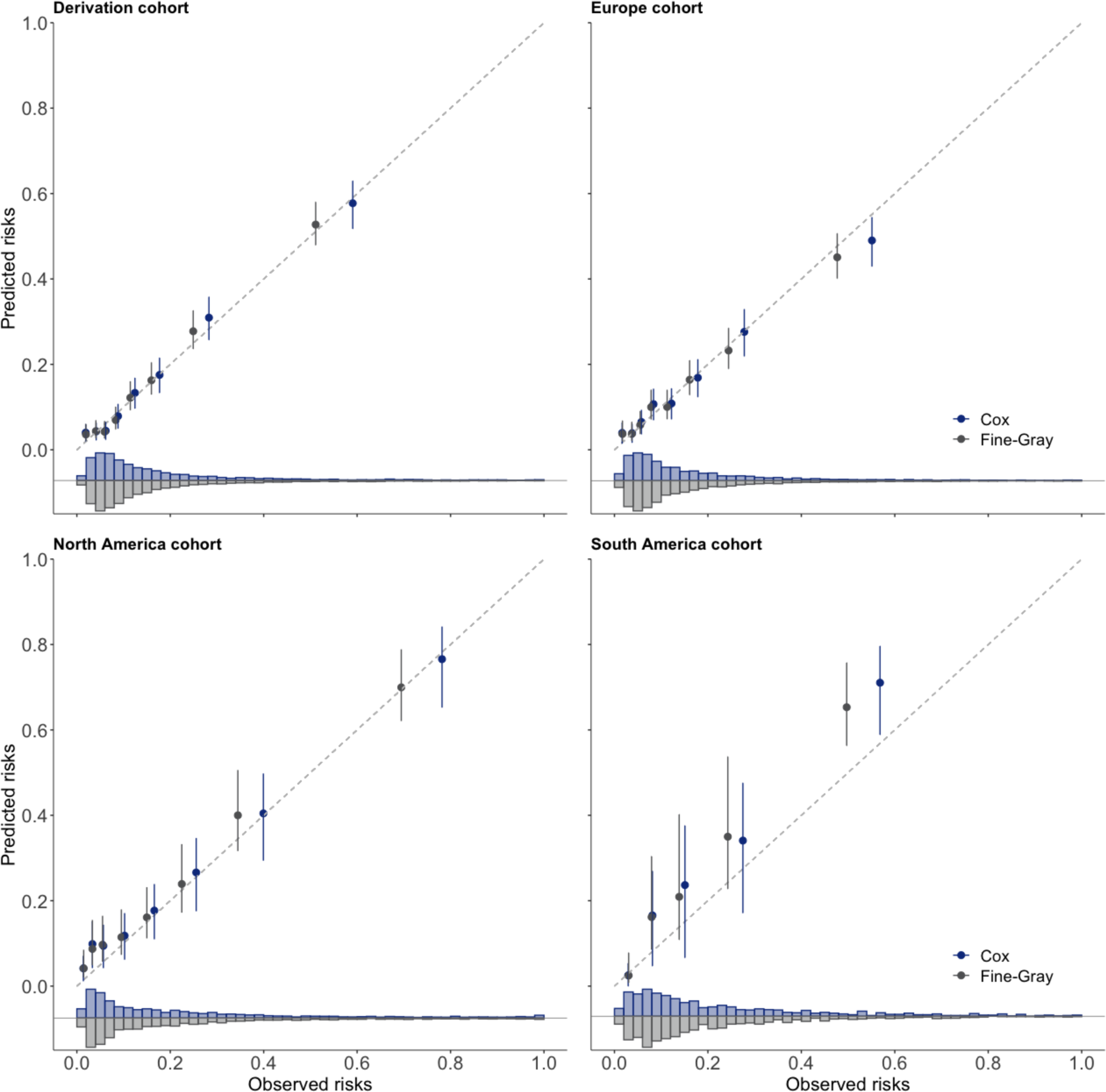
Calibration curves assessed at 7 years post risk-evaluation in the derivation and external validation cohorts. Calibration plots are presented at 7 years post risk-evaluation in the derivation cohort and in the European and North American external validation cohorts, and at 5 years post-risk evaluation in the South American external validation cohort. In each group (8 groups for the derivation, European and North American cohorts, and 5 groups for the South American cohort), the median of the predicted risks (one minus the individual predicted survival probabilities for the Cox model and the individual predicted risks, i.e., cumulative incidence, for the Fine-Gray model) was plotted against the observed event probability estimated by (one minus) the Kaplan-Meier estimator for the Cox model and the Aalen-Johansen estimator for the Fine-Gray model, respectively. The diagonal line at the origin represents the perfectly calibrated model. The histograms represent the distribution of the Cox and Fine-Gray models’ individual predicted risks.

Comparison of the relative calibration of the two prediction models with the Integrated Calibration Index, E50, E90, Emax and the root squared bias are presented in supplementary table 3. Both models exhibited close values in the derivation, North American and South American cohorts, with slightly smaller (better) metrics for the Cox model, whereas in the European cohort smaller metrics were obtained with the Fine-Gray model.

#### Overall fit

Across all cohorts, differences were minimal between the Brier Scores and the Integrated Brier Scores (IBS) of the two models. The IBS values amounted to 0.054 [CI 0.049;0.059] and 0.054 [CI 0.050;0.059] in the derivation cohort, 0.057 [CI 0.051;0.062] and 0.054 [CI 0.050;0.059] in the European cohort, 0.069 [CI 0.062;0.076] and 0.070 [CI 0.063;0.077] in the North American cohort and 0.076 [CI 0.064;0.088] and 0.079 [CI 0.067;0.090] in the South American cohort, for Cox and Fine-Gray, respectively. Differences were larger for IPA values, along with wider confidence intervals (table 2). Explained variation slightly differed in the Cox and Fine-Gray models in the derivation and North American cohorts, and was similar in the two other external validation cohorts.

### Subgroup analyses

We investigated the prediction performance of the models when applied in a series of distinct subpopulations in the derivation cohort, including living and deceased donors, according to donor and recipient age, recipient’s BMI, sex, and race, in highly sensitized and non-highly sensitized recipients, and in patients receiving induction by anti-interleukin-2 receptor or anti-thymocyte globulin (table 3). Overall, we found very good discriminative ability in all the subpopulations for both models. Discrimination, calibration and overall fit summaries were comparable between Cox and Fine-Gray model, and overall slightly better for the Cox model.

**Table 3.**
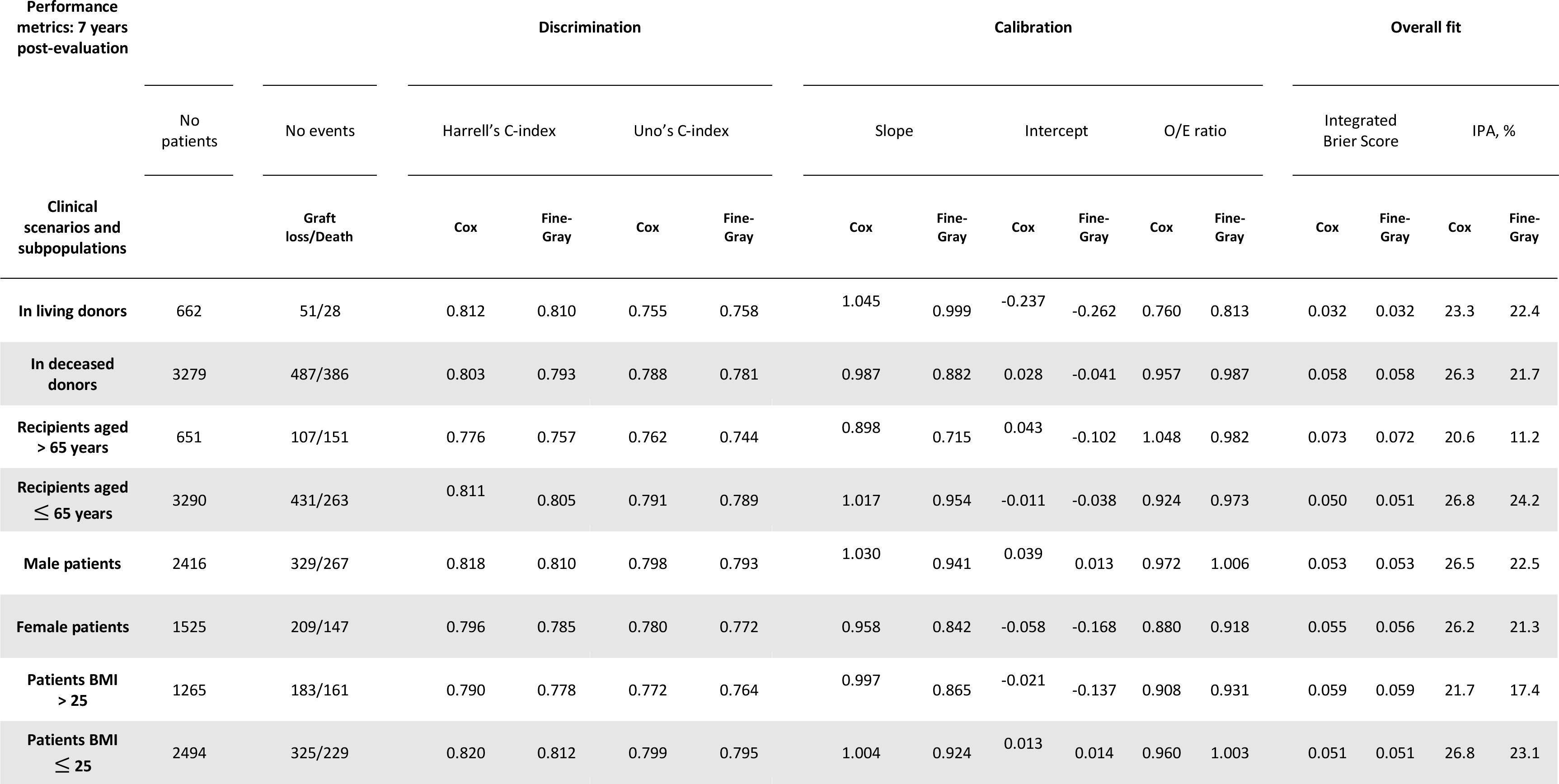
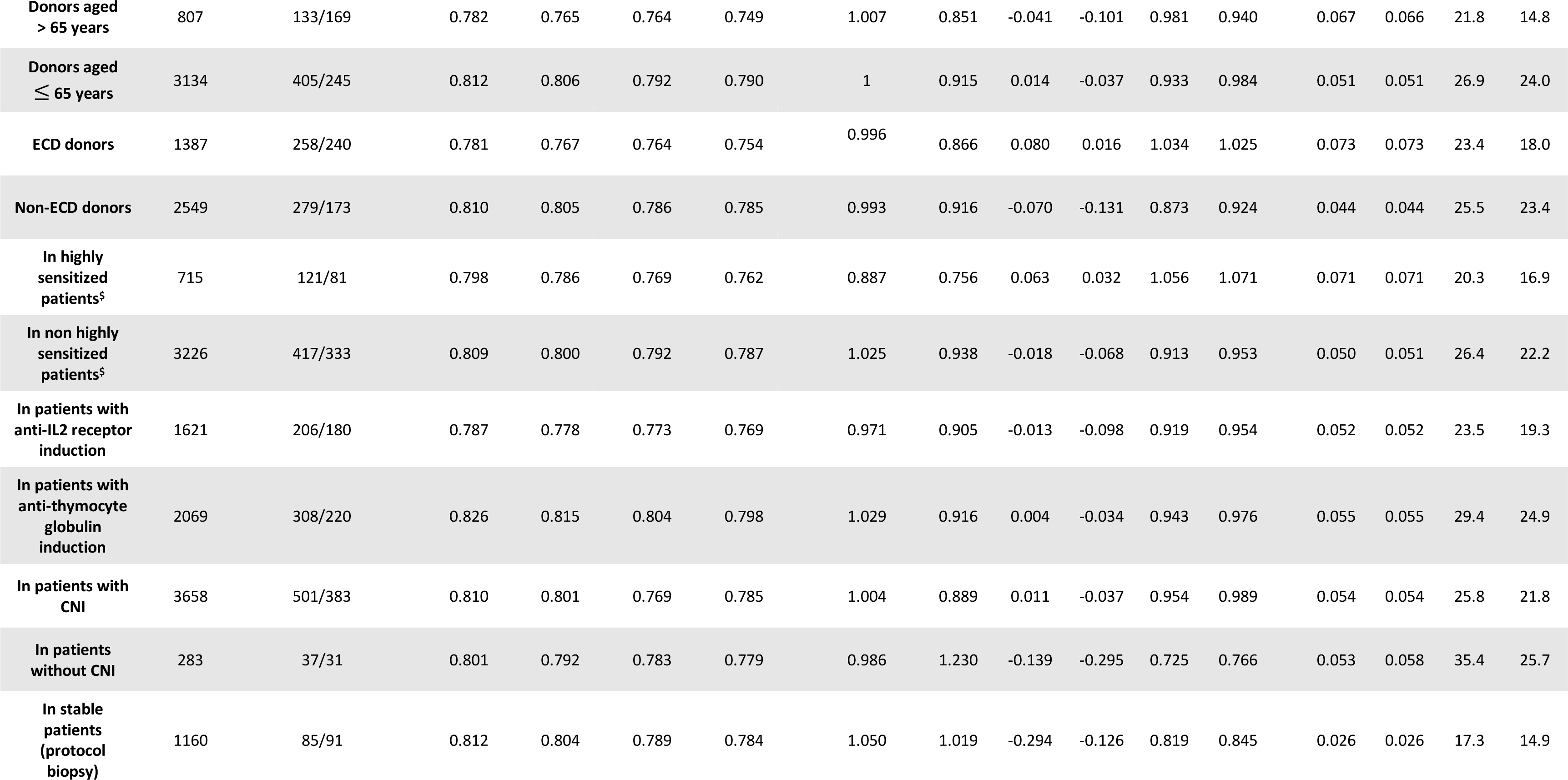
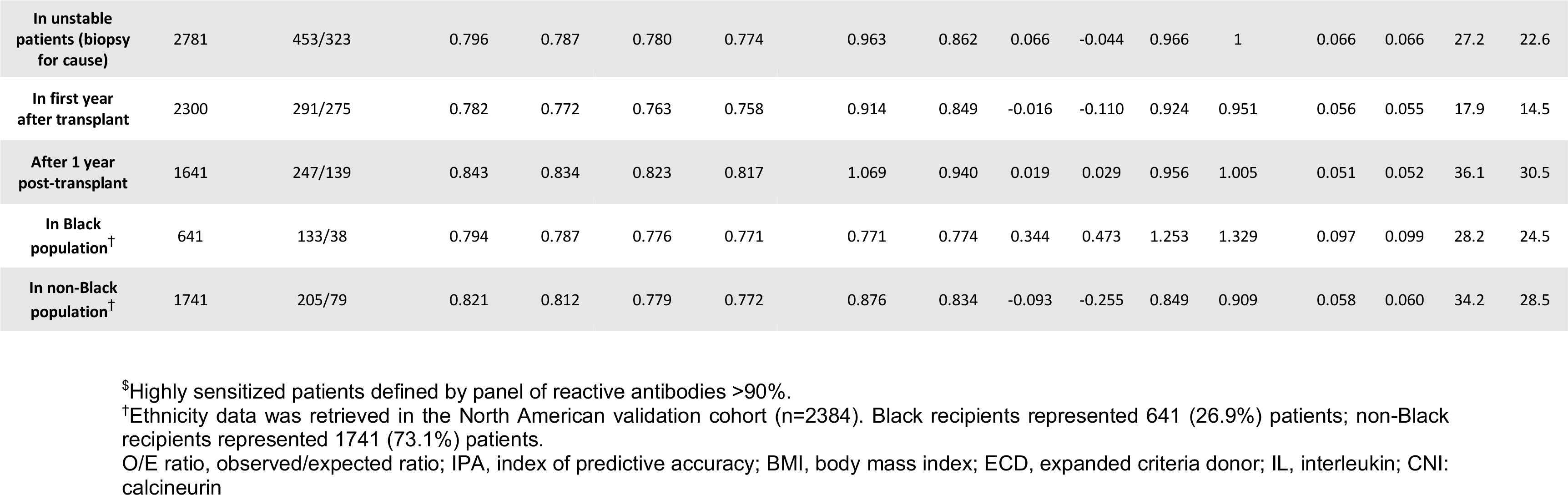
Predictive performance of the Cox and Fine-Gray models when assessed in different subpopulations and clinical scenarios in the derivation cohort at 7 years post risk-evaluation. Discrimination, calibration and overall accuracy performance metrics for the Cox and Fine-Gray models assessed at 7 years post risk-evaluation in the derivation cohort.

## DISCUSSION

### Overview

In this international study comprising 10 546 kidney transplant recipients, we developed prediction models for long-term kidney graft failure, with and without accounting for the competing risk of death. We performed a thorough assessment of these two models and showed consistency of allograft failure determinants, similar stability and comparable predictive performance across all validation cohorts. The findings were also consistent in a large series of subpopulations and clinical scenarios. To our knowledge, this is the largest study investigating in a comprehensive manner predictive performance in a competing and non-competing risks framework in large international prospective cohorts of kidney transplant recipients.

### Rate of competing events and impact on performance

In our study, the rates of death with a functioning graft were lower than the rates of graft loss in the derivation and validation cohorts. The absolute biases were relatively small in the overall cohort. In this setting, censoring death with a functioning graft or accounting for this competing event in the modelling strategy resulted in similar predictive accuracy. Similarly, a recent study by Clift *et al.*^48^ showed accurate and comparable discrimination and calibration of a standard Cox regression model and a competing risks regression model for long-term breast cancer related mortality prediction, in a large cohort where the event of interest was more prevalent than the competing event.

However, considering competing events may be crucial for cumulative incidence estimation and model performance when the amount of competing event is similar or higher to the amount of the event of interest, or when considering frail populations where patients are at higher risk of death, such as older recipients, or with comorbidities (e.g., cardiovascular diseases, hypertension, diabetes). This is reflected in our European validation cohort where the Fine-Gray model was slightly better calibrated than the Cox model.

For instance, in non-transplanted patients with severe chronic kidney disease, where the competing event of death is more frequent, several studies have shown that the overestimation of the cumulative incidence due to competing risk censoring increased with time^49,50^, and that a Fine-Gray model for kidney replacement therapy prediction achieved better discrimination and calibration than a standard Cox model^51^. Nevertheless, in our derivation cohort of kidney transplant recipients, performance in subpopulations including older donors and recipients, and in many other clinical scenarios, remained comparable.

### Prognostication and competing risks

In nephrology research, as well as in other medical specialties, there has been a growing call over the past years for the integration of competing risks analysis into prognostic modelling^7,52,18,19^. In contrast, for etiological purposes, hazard ratios from Cox models should remain the preferred approach to explore the associations between risk factors and the outcome^53^. In prognostic research, predictive accuracy and generalizability should remain the final judgment criterion for the benefit of the patient. If competing events are infrequent, a prediction model that does not account for competing risks may still accurately reflect the absolute risk for the population from which it was derived. However, if the model is validated in other populations where the competing events are more frequent, predictive performances may be impacted, although this impact may not be considered clinically significant and has to be demonstrated. Therefore, the modelling approach must be chosen based on the study design, the research question, the target populations, and the incidence of the competing event.

### Validation should rely on discrimination, calibration, overall fit

Validation of these prediction models should rely on an extensive evaluation of their discrimination, calibration, and overall fit. A single measure of discrimination and calibration does not offer a comprehensive view and is not sufficient to draw conclusions about an improvement in performance. This, nonetheless, is a general recommendation that holds true regardless of the modelling framework, and has been recently re-emphasized^6^. In the present study, a total of 15 metrics were used to provide a full picture of Cox and Fine-Gray models’ predictions, and showed consistency and stability of their predictions, and comparable prediction performance.

### Facilitating the implementation of competing risks analysis

An extensive literature exists on how to assess performance of prediction models in the absence of competing risks^4,5,6^. When handling competing risks, although statistical tools are well-known, comprehensive guidance for the evaluation of the same prediction performance has only been recently extensively addressed by Van Geloven *et al*^23^. Beyond this, calibration and discrimination metrics adapted for competing risk settings are scattered across original studies or methodological papers. For most of them, their implementation is often less straightforward and require more processing such as using pseudo-observations. There is therefore a contrast between the high number of studies recommending the use of competing risk models, and relatively few papers proposing an analytical framework to facilitate their implementation in research. The present study also aimed to contribute to fill that gap.

### Competing risks and machine learning

Further complexity may arise when comparing prediction performance of non-regression models in a competing and non-competing risks setting. Several machine learning survival models have been adapted to handle the presence of competing risks, such as random forests^54^ or neural networks^55,56,57,58^. However, the literature is still limited. Depending on the nature of machine learning models’ predictions, comparing their discrimination and calibration performances may be less straightforward since it typically requires further prediction transformations. Recent studies suggest that in low-dimensional settings, competing risks machine learning models provide similar discrimination but show miscalibration compared to competing risks regression models^48,59^.

### The advantages of deeply phenotyped cohort compared with registries

One strength of our study is the use of a large, unselected, prospective, deeply phenotyped multicentric cohort of kidney transplant recipients, which comprises key candidate risk factors for prognostic research, such as clinical, functional, immunologic, and histologic parameters. Using a cohort specifically designed for risk prediction represents an advantage, compared to the use of data coming from registries. Registry data may suffer from low quality, including lack of complete patient phenotyping, missing data candidate risk factors, long-term missing registered deaths or graft losses, and lack of follow-up and updates at fixed time points^60^. These intrinsic shortcomings may prevent them from being fit for purpose for making long-term predictions, thus limiting their value for prognostic studies.

### Limitations

This study has limitations. Firstly, we did not investigate other competing risk approaches, such as cause-specific Cox or pseudo-observations regression^61^. We focused our analyses on the Fine-Gray model, due to its wide use in medical literature for the analysis of time-to-event outcomes in the presence of competing risks.

Secondly, we did not investigate the clinical utility of these models, using metrics such as decision curve and net benefit. We preferred to focus on measures of discrimination, calibration, and overall fit, as these are the most crucial steps to evaluate a model before considering its clinical usefulness.

Thirdly, our study focuses on kidney transplant recipients with a low rate of death with a functioning graft. Our conclusions might not be generalized to other medical specialties or other populations, especially frail individuals highly susceptible to competing risks.

### Conclusion

Our study showed in a large, deeply phenotyped population of kidney transplant recipients with a low rate of death with a functioning graft, that a competing and non-competing risks model performed similarly in predicting long-term kidney graft failure. This is not to be interpreted as a general conclusion for competing risks modelling. Depending on the clinical scenario and the population considered, competing risks may be crucial to considerer and, consequently, competing risks models can contribute to more accurate prediction of graft failure.

## Data Availability

All data produced in the present study are available upon reasonable request to the authors

